# Defining movement instabilities in yips golfers using motion capture and muscle synergies

**DOI:** 10.1101/2020.08.19.20178475

**Authors:** Gajanan S. Revankar, Issei Ogasawara, Noriaki Hattori, Yuta Kajiyama, Shingo Shimoda, Alvaro Costa Garcia, Yuki Uno, Tomohito Nakano, Yasufumi Gon, Sadahito Kawamura, Ken Nakata, Hideki Mochizuki

## Abstract

‘Yips’ is an involuntary movement disorder seen in some professional golfers. The diagnostic challenge in yips is to distinguish symptoms of task-specific dystonia from psychological ‘choking’. We evaluated 15 professional golfers with mild symptomatic yips via anxiety tests, motion-capture and surface electromyography during a ‘putting’ task. Movement instabilities were analyzed via temporal statistical methodologies (one-dimensional statistical parametric mapping). In a subset of golfers, we found significant differences in angular velocities of the putter-club rotation and altered synergy neural coefficients during the downswing phase. Our results showed that golfers with mild yips require sensitive motion-capture evaluations wherein movement instabilities become evident. Particularly the downswing is affected, and the ensuing perturbations in phasic muscle activity share dystonic features that are consistently identified as abnormal muscle synergy patterns. Despite a strong subjective feeling of yips that golfers complain of, movement analysis can reliably exclude those with ‘choking’ from those with task-specific dystonias.

## Introduction

Some golfers experience an acute involuntary loss of performance in competitive environments. Instinctively, it is attributed to competition stress or anxiety that is seen in any high-precision, high-pressure sports. This performance deficit is classically known as ‘choking’^1,2^, and among golfers well- known as ‘the yips’. However yips is also suggested to be movement disorder. As a motor problem, it is characterized by abnormal involuntary twitching, jerks, spasms or freezing of planned motor movement^3^. Consequently, many authors regard yips as a task-specific dystonia^4–6^ or an occupational cramp resulting from intensive over-use of specific musculature for long periods of time which affects fine motor control^7^.

A key challenge in yips is to disentangle features of dystonia from ‘choking’^4,5^. Like all movement disorders, diagnosing task-specific dystonia is clinical. Neurologists have to ‘see it to diagnose it’ via physical examination or by video evidence ^8^, or risk categorizing yips as a psychological ‘choking’ phenomenon. To identify dystonic features in yips, it is crucial to observe or evoke patterns such as abnormal co-contractions during the task^9^ (e.g. in putting or drive shots). Unfortunately dystonic symptoms are rarely observed in golfers until their performance gets worse^10^. When symptoms of yips are mild, the movement variability is often inconsistent^11^ and detection of motor deficits may be difficult to capture videographically. Furthermore, dystonic patterns such as mild tremoric representations are dampened due to a two-handed club grip, and single handed shots to evoke yips may appear contrived^12^.

Therefore, our aim in this study was to objectively identify features of task-specific dystonia to rule-out ‘choking’ in mild yips golfers. The fallout of ‘choking’ or dystonia is singular – a measurable kinematic outcome with demonstrable performance loss due to an abnormal motor or stress response. We speculate that despite the strong psychological subjective priors associated with yips^1^, precise movement analysis would allow us to capture true dystonic features consistently in the form of abnormal stereotyped muscle activity. To that end, we designed our study around golfers suffering from putter’s yips, since putting, by virtue of its importance in competitive golf, is one of the most affected stroke in yips^13^. We evaluated ‘choking’ characteristics using trait and situational anxiety tests. To address the kinematics of putting shots, we used sensitive motion-capture systems to visualize movement characteristics and study their putting trajectories. Motion capture provides a direct image of orientation and position of the golf club during a putting stroke making it an ideal tool to quantify movement instabilities in golfers^14,15^. Within the framework of dynamic motor control, we applied the concept of muscle synergies on surface electromyographic recordings to represent the coordinated activation of muscle groups working as a specialized functional unit^16^. Prior studies have used synergy estimation as a tool to evaluate functional abnormalities between healthy and neurological patients^17–19^. Typically, muscle synergies function to access the best subset from a vast library of motor tasks to accomplish a smooth coordinated movement^20^. However in diseases of the nervous system such as stroke, dystonia or spinal cord injury, these physiological synergies are affected^18,19,21^. In functional movement disorders like task-specific dystonia, it is suggested that subjects may have fixed and normal synergy structures but abnormal neural coefficients that may indicate an inability to access or modulate a well-defined motor behavior^22^.

To summarize, we hypothesized that during putting, within each golfer, the co-contraction balance maintained by upper-arm and forearm muscles are altered in yips shots when compared to normal shot patterns. This difference would be observable via motion-tracking of the putter club together with muscle synergies to reveal features of movement instabilities in high-precision putting shots. These findings would allow us to characterize yips golfers with task-specific dystonia thereby ruling out those with choking.

## Methodology

### General information

15 professional golfers [14M, 1F, age 52.87 ±12.56 years (Mean, SD), all right handed] volunteered for the study and were prospectively enrolled at a single center. All golfers self-reported to have had yips symptoms for putting shots. Clinically we considered these symptoms as mild yips which were defined as inconsistencies during putting and those that were not strictly confined to problems only in putting^11,23^. Inclusion criteria were (i) professionals or high ranked golfers with current or past history of the yips, (ii) golfing experience of >15 years, (iii) handicap score of <14 before onset of yips and (iv) symptoms of yips severe enough to change grip style or alter training conditions. Exclusion criteria were subjects with (i) apparent physical injuries to upper arm or forearm (ii) diagnosed neurological, neuromuscular or psychological symptomatology. The research protocol was approved by the local ethics committee and written informed consent was obtained from the golfers in accordance to the Helsinki Declaration. All golfers were examined by a study neurologist on their arrival to the testing center.

### Anxiety tests and Putting task

Subjects were asked to complete two separate anxiety questionnaires at the start of the test session; (i) Trait Anxiety Inventory in Sports (TAIS) - which provides a comprehensive measure of anxiety in competitive sports^24^; and (ii) Sports Competition Anxiety (SCA) test - an index of situational anxiety that analyzes athletes on how they feel before and during a competitive situation^25,26^. The TAIS test uses a 4-point Likert scale for a set of 25 questions with a minimum score of 25 (low anxiety) and a maximum of 100 (high anxiety proneness). The SCA test consists of 15 items, 10 of which are scored, with a score of less than 17 indicating a low level of anxiety, 17 to 24 an average level of anxiety, and more than 24 a high level of anxiety.

The putting task was performed on an artificial putting surface, in a room equipped with 12 motion tracking cameras with the distance from starting position to hole set to 2.2 meters. 40 trials were performed and *after every shot*, the golfers were requested to verbally communicate their impression regarding their performance. The golfers were specifically instructed to try and putt *all* the shots without any explicit critique provided by the experimenter. Trials were then sorted and classified as normal-shots and yips-shots based on their subjective experience which was irrespective of their success in putting the shots. All trials were videotaped and qualitatively assessed for dystonic movements. Supplementary data provides additional details regarding evaluation of the putting task.

### Movement analysis using sEMG and motion capture

Fig. 1 provides the overview of surface electromyography (sEMG) and motion capture sensor evaluation. sEMG was recorded using Trigno wireless EMG system (Delsys, Inc. US) using 16 sEMG sensors sampled at 2kHz, band-pass filtered between 20 to 480Hz, rectified and low-pass filtered at 10Hz. Signals from biceps, triceps, pronator, supinator, flexor digitorum superficialis (FDS), extensor digitorum communis (EDC), extensor carpi radialis (ECR) and flexor carpi ulnaris (FCU) from each arm were recorded on LabChart-7 software (ADInstruments, New South Wales, Australia). A 12 camera OptiTrack Prime 17W system (NaturalPoint, Inc, US) was used for motion capture. The system was optimized for recording in small spaces with a high resolution of 1.7 Megapixel recorded at a frame rate of 360 frames per second.

**Fig. 1.**
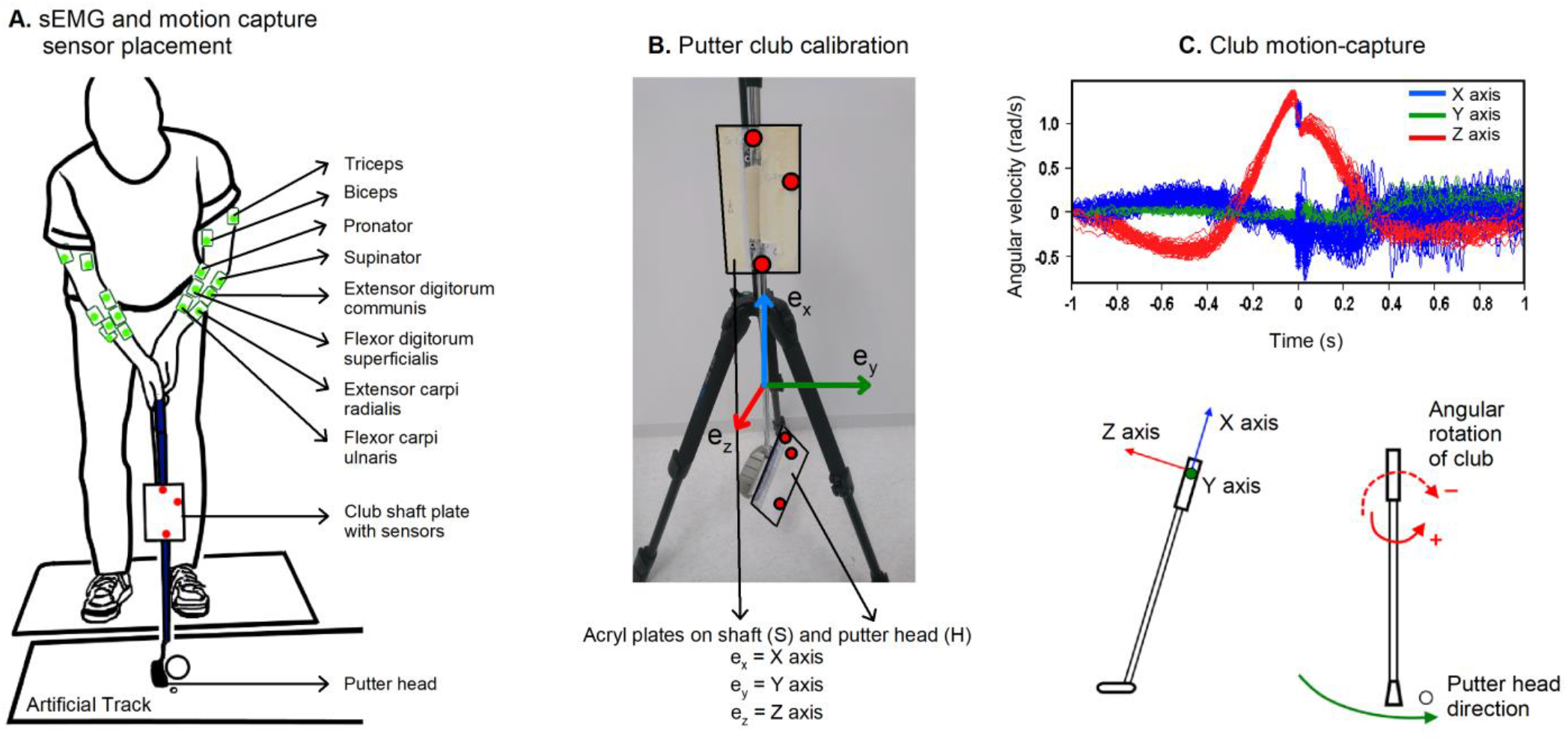
(A) Graphical representation of wireless sEMG electrodes used along with the sensors on the putter club for motion capture system, (B) The club coordinate system consisted of three orthogonal unit vectors ***e_x_***, ***e_y_***, and ***e_z_*** for X, Y and Z axis respectively which were calculated using acryl plates attached to shaft and putter- head. (C) (top) Snapshot of putting swing for an individual participant. Time ‘0’ = time of ball impact. (bottom) The club coordinate system were defined as ***e_y_*** – the unit vector in the direction of forward swing (Y axis), ***e_x_*** - the unit vector parallel to the club shaft (X axis) and ***e_z_*** - the vector perpendicular to ***e_x_*** and ***e_y_*** (Z axis). The angular velocity of putter head (red curve - Z axis) was used since this was the most sensitive parameter to record twitches, jerks or freezing of movements.

For each putting trial, motion tracking and sEMG data were epoched for 2 seconds (−1 second to +1 second with 0 being the time of ball impact). This epoch included a time window immediately prior to start of backswing, backswing, downswing till ball impact and follow-through phases. The club coordinate system was defined to monitor the angular velocities of the club during the putting stroke. Calculations of angular velocities are described in supplementary data. The ‘putter downswing’ is a fast movement occurring at a critical time point between backswing (governed by anti-gravity muscles) and follow-through (by muscles defining posture stability necessary to end the swing phase). Since the downswing phase in the golfers depended on the putter head direction and speed, we focused our analysis on the phasic component of the putter swing^20^. Given that a constant tone was involved to maintain the stability of the putting movement, the phasic component was extracted by subtracting the tonic component represented as a linear ramp between 400ms before movement onset (backswing) and 400ms after follow-through phase^20^.

For muscle synergy analysis, trials were downsampled from 2 kHz to 500 Hz and synergies were extracted for the downswing phase from each arm separately. Phasic sEMG data described above, for every trial, was factorized using non-negative matrix factorization (NNMF) into a synergy matrix of weights (W), a neural recruitment coefficient (C) represented mathematically as:

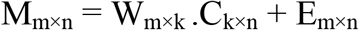

where M is a *m×n* matrix of sEMG data (with *m* = number of muscles and *n* = number of samples), W is a *m×k* matrix containing the muscle synergies used to reduce *m* muscles into *k* dimensional space, and C is a *k×n* matrix containing the *k* patterns used to control full set of *m* muscles temporally defined by *n*. *k* denotes the number of synergies and E the residuals. Intuitively, for a particular muscle activation pattern M, W specifies the relative contributions of the muscles involved in the synergy and C is the coefficient that changes over time and across conditions. To avoid the local minima, the algorithm was iterated 1000 times and the final synergy vectors were normalised by their maximum values^27^. The methodology for calculation, validation of synergies and their reconstructions were performed as described in previous studies^28,29^. The NNMF algorithm received each trial data as input and synergies were calculated trial by trial without fixing the synergy number. The least synergy number to adequately reconstruct the sEMG data were quantified by 2 parameters - the centered Pearson’s correlation coefficient (R^2^) calculated for every muscle from the trial dataset with respect to the reconstructed synergies and the variance accounted for (VAF) which is used to obtain a goodness of fit between the actual and reconstructed EMG from the synergies. To obtain consistent features from the data, such of those synergies which crossed a threshold of mean value R^2^ (EMG_reconstructed_ R^2^) of 80%, a VAF of 90% and a VAF of muscles of 80% were considered sufficient to represent the input EMG dataset. In order to match the resulting synergies, we sorted the trial based synergies based on their degree of alignment represented by their Pearson’s correlation coefficients and matched them versus every pair. As a first step, synergies from a trial (any) within the group (normal or yips) were sorted according to their power contribution to the filtered sEMG data. sEMG power was computed as the Root Mean Squares (RMS) of the sEMG signals recorded. In the second step, the synergies from the rest of the trials were ordered with respect to the correlation of their weights obtained from the previous step.

### Statistical analysis

Descriptive and correlational statistics for relevant demographic variables were performed. We identified the total number of yips trials and then a random number of normal-shot trials were matched to keep the number same under all response conditions. To evaluate the motion tracking differences between normal and yips shots, a 2-tailed paired t-test using 1-dimensional Statistical Parametric Mapping (SPM) was used. The procedure involved calculating a t-statistic threshold (t*) and the temporal smoothness at each time point using the residuals on the time-series data^30^. For each subject, thresholds were calculated based on the number of matched trials which differed for each participant making the SPM t-test ideal for subject-level analysis. Alpha value of 0.05 was set, and if the SPM t-trajectory crossed the threshold any time point in the time series data, the values were deemed significant (Fig.2A, Comparison)^31^. The advantage of this method is that the results are reproducible and avoids dependence on standard deviations to make interpretations where sEMG often show variability due to multiple trials. For the resulting synergy weights, paired t-tests with Bonferroni correction were applied, and for the temporal neural coefficients a 2-tailed paired SPM t-test was used (Fig.2B, Comparison). For both tests, statistical significance was set to p<0.05. All offline analyses were done using Matlab 2017b. SPM analyses were performed using open-source spm1d code available at www.spm1d.org.

#### Data sharing statement

Data that support the findings of this study are available from the corresponding author upon reasonable request. Custom scripts will be available before publication and deposited in a community repository.

**Fig. 2.**
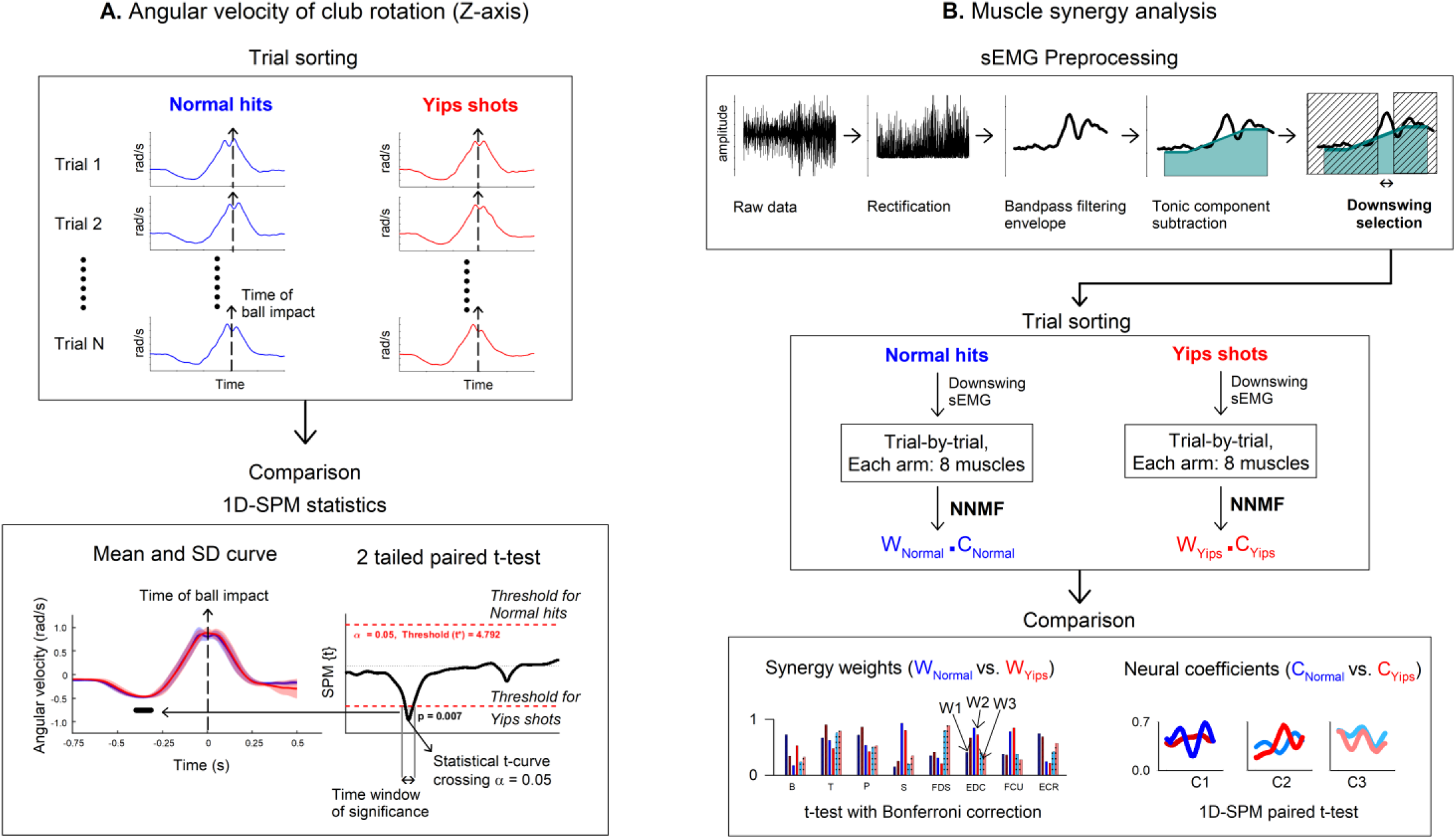
Graphical representation of preprocessing procedures and statistical methods used in the study. (A) Analysis of angular velocity of club: For each subject, trials were sorted to perform comparisons using one dimensional statistical parametric mapping (1D-SPM). For normal hits and yips shots, the mean value of angular velocity curves, trial-by-trial, from backswing to impact and follow-through were mostly similar (overlapping mean SD curves) except for certain time segments during downswing (shown by solid black line in mean SD curve). This represents the time where the values for yips shot were higher than for normal hits crossing a critical threshold (t*) in paired SPM t-test. The probability of finding such significant time segments by random sampling is given by their respective p values < 0.05 for individual subject thus rejecting the null hypothesis. (B) Muscle synergy analysis: sEMG (surface electromyography) trials were first preprocessed to obtain phasic downswing component (preprocessing steps shown here is for a single muscle). After trial sorting, for each arm comprising of 8 muscles each, synergy analysis was performed using NNMF (non-negative matrix factorization) algorithm to obtain synergy weights (W) and neural coefficients (C) for normal and yips-shots respectively. Statistical comparisons were performed between normal and yips shots when reconstructions were > 80%, shown here for 3 synergies as W1, W2 and W3 for weights and C1, C2 and C3 for coefficients respectively. Legend: biceps (B), triceps (T), pronator (P), supinator (S), flexor digitorum superficialis (FDS), extensor digitorum communis (EDC), extensor carpi radialis (ECR) and flexor carpi ulnaris (FCU).

## Results

### Demographic details

Demographic details are described in Table 1. Participants in our study had long golfing experience [35 years, (26.2, 40.7) (Median, 1^st^ quartile, 3^rd^ quartile)], with considerable duration of yips symptoms [10 years (4.25, 23.2)]. Linear regression showed moderate significant correlation between playing experience and onset of yips symptoms (N=15, R^2^ =0.33, p=0.024). Pearson’s correlation coefficient for anxiety tests (TAIS score vs. SCA test) was statistically significant (N=15, R=0.73, p=0.002). None of the golfers showed video-graphic evidence of freezing, twitching or jerks of the upper arm or forearm during the task.

**Table 1.** TAIS score - Trait Anxiety Inventory in Sports score, SCA test - Sports Competition Anxiety test

| Golfers ID | Age | Golfing experience (years) | Symptom duration (years) | Response during experiment for yips shots classification | Anxiety tests |  |
| --- | --- | --- | --- | --- | --- | --- |
|  |  |  |  |  | TAIS score | SCA test |
| Subject 01 | 53 | 35 | 5 | "Arm freezes, doesn't appear to move smoothly" | 44 | 22 |
| Subject 02 | 48 | 30 | 4 | "Tensed, bit stressed, club movement feels slightly sticky" | 50 | 20 |
| Subject 03 | 54 | 36 | 10 | "Club feels like it rotates to the same side, like a twitch" | 42 | 19 |
| Subject 04 | 37 | 18 | 3 | "Stressed, arm feels cramped, gripped too tight, movement not smooth" | 45 | 27 |
| Subject 05 | 73 | 40 | 5 | "Feeling of ball not being hit where I want it to" | 34 | 18 |
| Subject 06 | 45 | 30 | 10 | "Movement not smooth, getting stressed when focused for too long" | 66 | 27 |
| Subject 07 | 84 | 45 | 25 | "Shots are too strong, too fast sometimes, grip and arm feels tight" | 52 | 25 |
| Subject 08 | 53 | 41 | 28 | "Slightly cramped in arm during backswing" | 49 | 14 |
| Subject 09 | 47 | 23 | 4 | "Arms feel cramped, tightened, stressed when focused for long." | 37 | 20 |
| Subject 10 | 59 | 36 | 31 | "Shots feel shifted involuntary, arm feels cramped and tensed, shots too strong" | 27 | 11 |
| Subject 11 | 38 | 25 | 2 | "Shots not smooth, downswing feels stuck" | 51 | 22 |
| Subject 12 | 41 | 31 | 15 | "Shots too strong, arms feels rigid and tensed" | 56 | 24 |
| Subject 13 | 59 | 42 | 8 | "Arm appears to freeze, shots appear shifted, tension in arms during downswing" | 61 | 25 |
| Subject 14 | 48 | 24 | 18 | "Downswing not smooth, twitches during downswing in both hands" | 60 | 25 |
| Subject 15 | 54 | 48 | 28 | "Shots too strong, grip intentionally strong, forearm tightened" | 54 | 22 |

### Motion tracking results

For the putting task, shots were classified as Normal-shots (Ns) and Yips-shots (Ys) according to their subjective responses as described in Table 1. About 25% of the shots were Ys (10.0 ±3.99) out of 40 trials. Quantitatively all participants showed some degree of overlap in swing trajectories (Mean, SD curves) between Ns and Ys, suggesting that high precision shots requiring fine control do not deviate much from the mean (Fig.3). During the swing phase, SPM paired t-test showed specific time segments of t-curves which crossed the critical threshold value at p < 0.05 - in 9 out of 15 golfers as shown in Table 2. This time window of significant change in angular velocity of the putter club between Ns and Ys was characteristically observed in the downswing phase. However the downswing time in itself remained similar between Ns and Ys within the subjects [Ns = 315.26 ms ±54.5, Ys = 315.87 ms ±55.9, paired t-test - t(14) = -0.49, p = 0.631] (Supplementary Table-1).

**Fig. 3.**
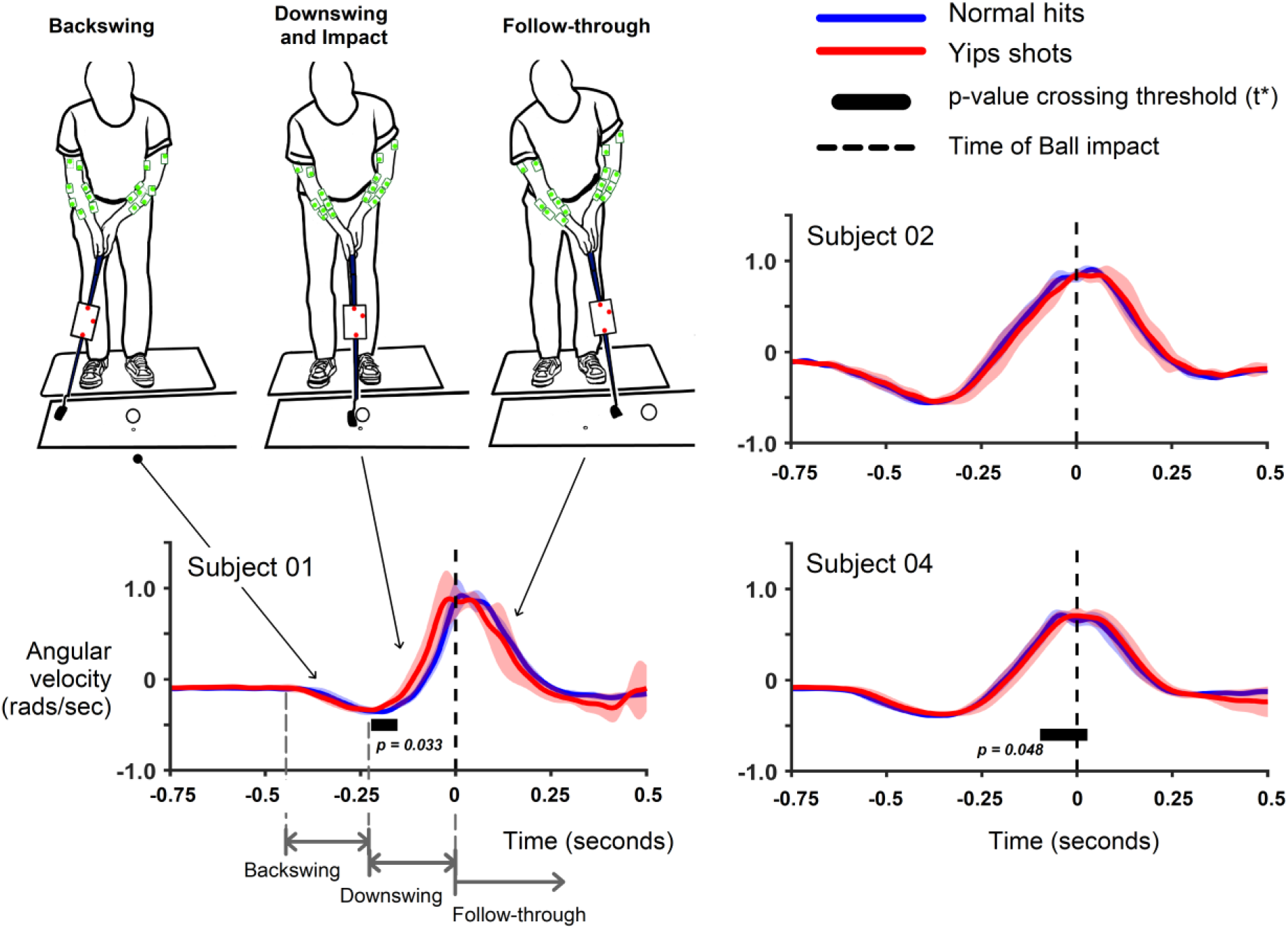
Mean and SD curves for angular velocity of the putter club in 3 example subjects - Subject 01 (number of matched trials, N=8), Subject 02 (N=7) and Subject 04 (N=13). Blue trace = Normal shots, Red trace = Yips shots. Sections of backswing, downswing and follow-through are shown in Subject 01 in grey dotted line with the black dotted line representing time of ball impact. For normal hits and yips shots, the mean value of angular velocity curves from backswing to impact and follow-through were mostly similar throughout except for certain time segments during downswing (shown by horizontal solid black line). This represents the time where the values for yips shot were higher than for normal hits crossing a critical threshold (t*) in 2-tailed paired SPM t-test.

### Synergy analysis results

Averaged sEMG between Ns and Ys failed to show any significant difference using mean and SD plots for each subject (Supplementary Fig.2) to evaluate for stereotyped burst patterns. Based on the changes observed in angular velocity, we pre-selected only the downswing time for muscle synergy analysis. Synergy constructions for short time periods have the advantage of revealing minor perturbations in the movement that affect stability^32^. Muscle synergies calculated iteratively for the downswing phase was represented by 3 synergies for each arm with reconstruction scores of approx. 80% and above. Reconstruction scores are shown in Supplementary Table-2.

The patterns of 3 spatial synergies (W1, W2 and W3) extracted from each arm showed broad similarities between Ns and Ys for the downswing task (Supplementary Fig. 3). For the left arm, W1 showed a synchronized burst with an extension component mainly involving the supinator, EDC and ECR. W2 had flexors active with strong activations in pronators, FDS and FCU. W3 showed a non-specific activation in all muscle groups. In contrast, the right arm synergies showed an opposite spatial structure. W1 was mostly observed having a flexor component with biceps, pronators, FDS and FCU showing strong activations. W2 showed higher activations in Sup, FDS and ECR compared to other muscles. Finally W3 had both higher pronator and supinator activation. Subject-wise comparison of paired t-test showed no significant changes in W’s between Ns and Ys.

For evaluation of neural coefficients (C), SPM paired t-tests showed significant differences in 11 of the 15 golfers (Table 2). The change in neural coefficients with respect to synergy weight is shown as an example in 2 golfers in Fig. 4. Since the downswing phase is a fast movement action, we construe this change in C’s to have affected the entire downswing time of interest (and not a specific time segment within the downswing phase).

**Fig. 4.**
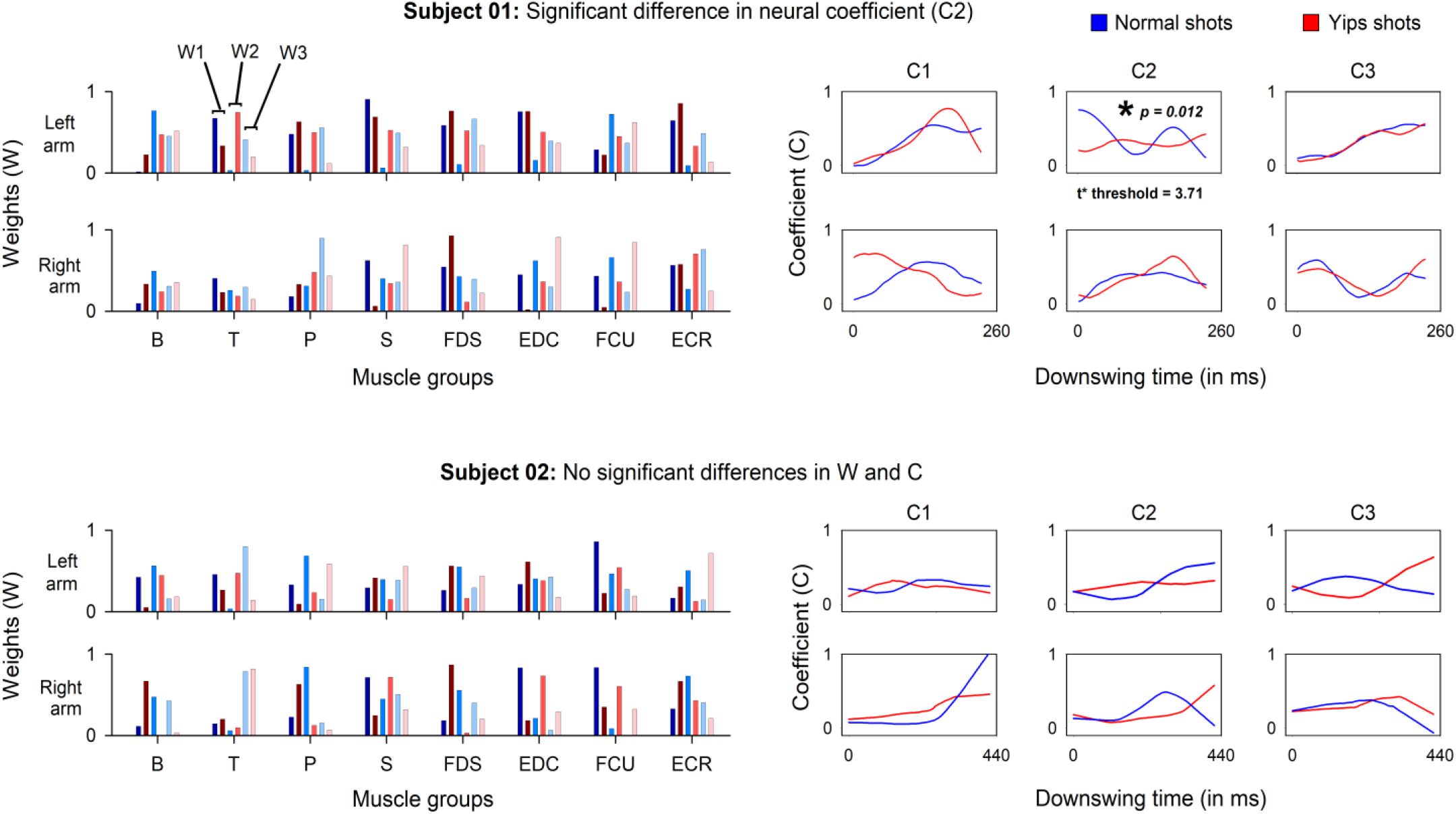
Example shows synergy decomposition for 2 golfers, Subject 01 (number of matched trials N=8) and Subject 02 (N=7). Three synergies for each arm were calculated. Mean muscle activation weights (W) are shown as W1, W2 and W3 in (shades of) blue for normal hits and in (shades of) red for yips shots. Similarly mean neural coefficients (C) are shown as C1, C2 and C3 for the 3 synergies respectively. Muscle groups shown are biceps (B), triceps (T), pronator (P), supinator (S), flexor digitorum superficialis (FDS), extensor digitorum communis (EDC), flexor carpi ulnaris (FCU) and extensor carpi radialis (ECR). While synergy weights were mostly similar subject-wise (paired t-test with Bonferroni correction), neural coefficients showed significant difference for Subject 01 between normal and yips shots (SPM 2-tailed paired t-test). No such changes were observed for Subject 02.

Compiling the findings on movement analysis, we observed that Subject 02, 07 and 15 did not show any differences between normal and yips shots on motion capture or on muscle synergy analysis. The remaining golfers showed significant differences in either motion capture (Subject 03) or muscle synergy analysis (Subject 06, 10, 12) or frequently in both (Table 2).

**Table 2.** SPM – 1-dimension Statistical Parametric Mapping, dF = Degrees of freedom, t* value – threshold set for alpha = 0.05, N.S – Not significant, ms = milliseconds.

| Golfer ID | Total yips trials | Angular velocity<br>(SPM paired t-test) |  |  |  | Muscle synergy<br>(Neural coefficients (C) - SPM paired t-test) |  |  |  |  |
| --- | --- | --- | --- | --- | --- | --- | --- | --- | --- | --- |
|  |  | *t\**<br>value | *p* value | *Affected time (pre-impact)* |  | *Affected arm* | *dF* | *Synergy number* | *t\**<br>value | *p* value |
| Subject 01 | 8 | 6.75 | **0.033** | 170 ms |  | Left | 1, 7 | 2 | 3.71 | **0.012** |
| Subject 02 | 7 | N.S | N.S | N.S |  | N.S | N.S | N.S | N.S | N.S |
| Subject 03 | 8 | 5.67 | **0.013** | 160 ms |  | N.S | N.S | N.S | N.S | N.S |
| Subject 04 | 13 | 4.33 | **0.048** | 80 ms |  | Right | 1, 12 | 1 | 3.34 | **0.009** |
| Subject 05 | 8 | 6.27 | **0.046** | 265 ms |  | Left | 1, 7 | 2 | 4.18 | **0.046** |
| Subject 06 | 8 | N.S | N.S | N.S |  | Left | 1, 7 | 2 | 3.90 | **0.041** |
| Subject 07 | 7 | N.S | N.S | N.S |  | N.S | N.S | N.S | N.S | N.S |
| Subject 08 | 10 | 5.13 | **0.024** | 175 ms |  | Left | 1, 9 | 2 | 3.38 | **0.019** |
| Subject 09 | 9 | 6.71 | **0.044** | 10 ms |  | Left | 1, 7 | 2 | 3.81 | **0.036** |
| Subject 10 | 22 | N.S | N.S | N.S |  | Right | 1, 17 | 2 | 2.94 | **0.034** |
| Subject 11 | 8 | 5.84 | **0.024** | 210 ms |  | Right | 1, 7 | 1 | 4.02 | **0.000** |
| Subject 12 | 8 | N.S | N.S | N.S |  | Right | 1, 7 | 1 | 3.79 | **0.000** |
| Subject 13 | 9 | 5.74 | **0.022** | 15 ms |  | Right | 1, 8 | 1 | 3.77 | **0.001** |
| Subject 14 | 11 | 4.89 | **0.010** | 190 ms |  | Left | 1, 10 | 2 | 3.37 | **0.026** |
| Subject 15 | 15 | N.S | N.S | N.S |  | N.S | N.S | N.S | N.S | N.S |

## Discussion

Our study is the first comprehensive report on mild yips golfers where sensitive movement-related measurements were utilized to reveal features of a movement disorder. Specifically we found that in mild yips (i) golfers experience reasonable amounts of stress that may contribute to a state of underperformance overlapping with their movement instabilities; (ii) for putting shots, whereas motion-tracking readily captures fine motor changes in movement trajectories, features of co-contraction imbalance on sEMG recordings may not be particularly evident; (iii) finally, the downswing is particularly affected, and the ensuing perturbations in muscle activity share dystonic features that are consistently identified as abnormal muscle synergy patterns.

### Stress and competitive sports

Models that explain sports related anxiety conceptualize that the cognitive self-evaluation and stress response if left unchecked, result in increased muscle tension, loss of focus and a range of other physiological behavioral changes^33^. As a consequence, depending on the individual’s own threshold of sense of anxiety, the performance-anxiety loop can either streamline their quality of shots or can potentially debilitate the task^34^. The golfers in our study were not of anxious type as revealed by TAIS scores but experienced a certain degree of competition stress as seen from SCA test. Though we believe this to be normal stress responses during gameplay, the subjective feedbacks given during the experiment suggest otherwise (Table-1). Qualitatively, there appeared to be less disagreement among our golfers that anxiety was perhaps not the only factor contributing to their performance deficit. Prior studies have also reported similar observations that higher muscle activations and grip force can impact stroke play kinematics irrespective of levels of situation induced anxiety^6,35,36^.

### Downswing putting accuracy in yips

Professional golfers frequently spend considerable time in perfecting the putting stroke^37^. To perform a smooth shot, expert golfers recommend that the start of downswing phase of the club to be dictated by gravity, then eventually adjusting the hand torque at ball impact. Of significance is the angular velocity of the putter club-head and the hand torque model which advocates minimizing the hand torque from the start of downswing to allow a less variable velocity at ball impact, making the putting shots more consistent and accurate^38^. As an outcome measure of motion patterns, we chose the putter-club angular velocity during the entire swing and found that it was largely inconsistent during the downswing phase for yips-shots. This result reflects the temporal difference between normal and yips-shots and hence indicates the change in the uniformity or regularity of the shots. We therefore interpret that the inconsistencies seen for yips-shots is a miscommunication in the co-contracting forearm muscles during such „fine adjustments’ that may have prevented an ideal trajectory anticipated by the golfers.

### Utility of muscle synergies

Our initial screening of sEMG differences in multiple muscle pairs between normal and yips-shots was mostly inconclusive (Supplementary Fig.2). Adler et. al. reported that abnormal co-contraction patterns were observed in wrist flexors and extensors in the downswing phase in yips affected golfers^9^. Co-contractions are essential to maintain the joint position balance and in high-precision shots like putting, any discrepancy that results in an erratic trajectory may not necessarily imply muscle dysfunction^39^. Therefore in low-force tasks like putting, due to the trial by trial sEMG variability, we abstained from over-reporting the effect of co-contractions as manifestations of yips in our participants.

This leads us to the next point in using synergy analysis to identify features of focal dystonia. In maintaining biomechanically constrained joint balance, we used muscle synergies to identify patterns of muscle activity that achieve multi-joint coordination. We observed that the muscles of the elbow-wrist joint required 3 synergies to provide the necessary balance, direction and speed to perform the putting stroke. The apparently high number of synergies for a putting stroke documented here is a response for a low force isometric task which necessitates precise movement control^40^.

The spatial synergy weights (W) represent the muscle activations during a specific time of interest, here, the entire course of the downswing. In maintaining downswing balance, we observed that the variability in W’s was generally seen to be constrained to similar spatial patterns for normal and yips-shots. This appears to be an expected outcome since expert golfers minimize movement at the wrists by locking them in position, control positional parameters by spatially scaling downswing times and orient club head to avoid change in trajectories^41^. Furthermore, the extracted W’s illustrate functional groupings and due to their anatomical proximity or effect of crosstalk, we speculate that this may have contributed to the similarity in spatial synergies.

Neural coefficients (C) are believed to represent neural commands from specific synergies that influences the W’s modulating it over time^28^. The observed differences in C’s in a subset of golfers signify an altered phasic muscle synergy activity in yips-shots than in normal conditions. These were uniquely defined for each golfer suggesting an individual-specific relationship in muscle activations from higher centers. Muscle activations which occur in a multi-dimension space, require a coordinative input in the form of neural information to exclude and select appropriate motor patterns to harmonize movement. This harmony is achieved by spinal pre-motor neurons which dynamically adjust activations from inhibitory and excitatory pre-motor neurons in conjunction with higher centers like sensorimotor cortex, basal ganglia and cerebellum^19,42,43^. With long years of practice and repeated use, these pre-motor neurons evolve to reduce variability and strengthen access to a specific synergy necessary for motor control^44^. Yips-shots are an extreme example of this creation of ‘specific synergy’ due to a highly sensitive pool of pre-motor neurons eventually leading to abnormal sensory integration^45^, impaired cortico-motor information processing or maladaptive plasticity^46^. In actively adjusting putting trajectory, these golfers were unable to maintain their co-contraction stability due to an abnormal synergy representation. The manifestations seen here of yips-shots are therefore an amplification of altered dynamic phasic activity that dystonia are a part of.

### Limitations

There are some limitations to this study. Each golfer plays with a certain degree of uniqueness and this subtle but diverse behavior in motion capture and sEMG led us to focus on a case-by-case basis. Testing in laboratory environments never brings out the same level of anxiety experienced by players as ‘sinking the putt’ in the green. Our goal was not to create a high-stress environment for the golfer but rather identify features of muscle and kinematic imbalance under any possible yips-like condition. We were careful to interpret our findings on muscle synergies which were based on changes in unidirectional downswing movement. Detailed modeling using joint kinematics along with truncal muscle synergy estimation for putting shots could be beneficial to address in the future. Furthermore, it would be advantageous if a standardized anxiety test was specifically tailored to yips since it the first-line assessment for any yips affected athlete.

In our formulation, we focused on identifying features of dystonia via movement analysis though other crucial variables may also be at play. Using demographic variables such as golfing experience, duration of yips symptoms, practice rounds per year, along with results from anxiety scores, motion-capture and synergy analysis, we categorized the participant groups into 2 types, using an unsupervised cluster analysis algorithm (Fig. 5 and Supplementary Table-3). The basis for this classification system comes from a frequently documented ‘continuum’ model suggested by Smith et.al. with Type-1 (dystonia) and Type-2 (choking) yips^4^. Non-movement associated variables could help classify the golfers better, although our focus rested mainly on motion-capture and muscle synergies to identify the problem. Future studies will need a systematic evaluation of these effects.

**Fig. 5.**
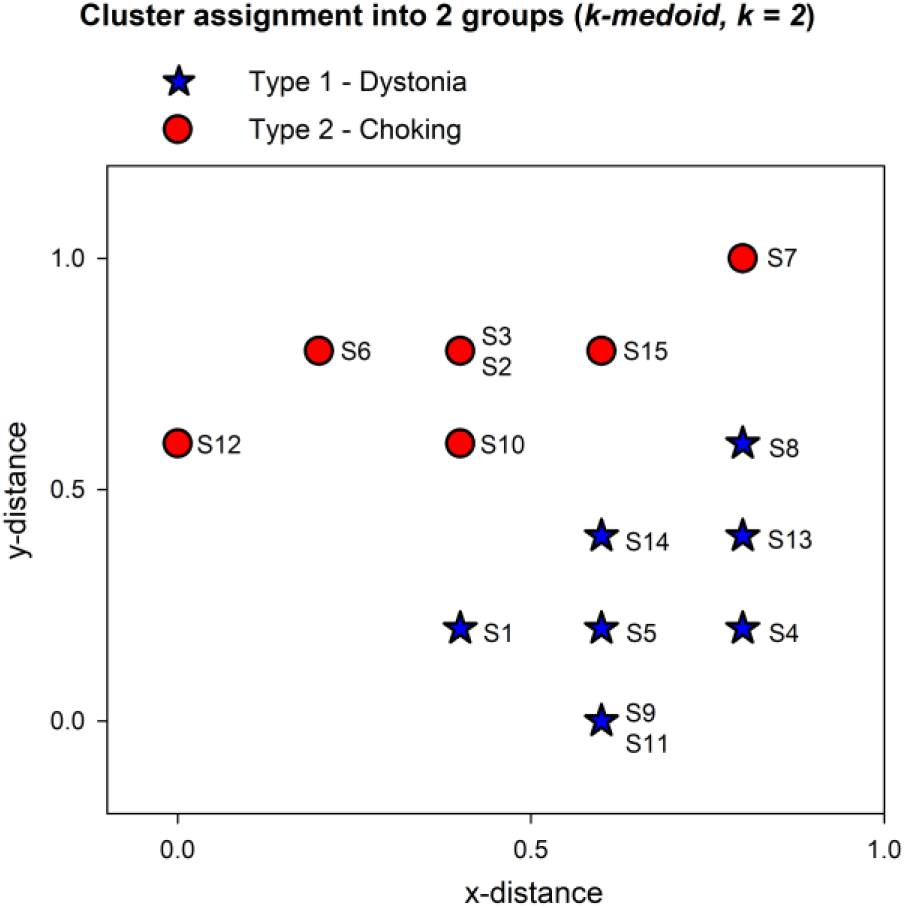
(A) Classification of yips golfers shown using two-dimensional scatterplot. The variable *k* for number of clusters was set to k = 2 to cluster the input data into Type-1 dystonia and Type-2 choking, suggested by Smith et. al. ^4^. The best total sum of distance was 5 for these two clusters.

## Conclusions

Diagnosis of yips is fraught with difficulties mainly due to limited research, scant literature and incongruity within the target populations. Still, we were able to highlight and unravel abnormal kinematics and synergy patterns that influence motor behavior among golfers irrespective of their subjective feeling of yips. Future work will need to address the link between spinal and central causes of yips, their mechanisms and how interventions could rehabilitate these golfers using behavioral therapy, swing dynamics or ‘normalize’ the faulty synergies leading to an improvement in their performance.

## Data Availability

Data that support the findings of this study are available from the corresponding author upon reasonable request.

## Acknowledgements

We express our sincere gratitude to the golfers who participated in this study, and special thanks to Kaho Umegaki, Megumi Nambo, Tei Kohten and Shinsuke Okubo who helped us in data collection.

## Author contributions

Conceptualization - G.S.R., I.O., N.H., Y.G., S.K., K.N. and H.M.

Methodology - G.S.R., I.O., Y.U., N.H., Y.K. and T.N.

Software, Validation - G.S.R., I.O., S.S. and A.C.G.

Formal analysis, Data curation, Writing – original draft preparation - G.S.R. and I.O.

Writing – review & editing - G.S.R., I.O., N.H., S.S., A.C.G., K.N. and H.M.

Supervision, Funding acquisition - K.N. and H.M.

## Subject-area

Neurology

